# Refining the biopsychosocial model of trauma: vulnerability and social support as primary predictors of mental disorders in a clinical sample

**DOI:** 10.64898/2026.05.25.26354043

**Authors:** Luciano Ferreira Rodrigues-Filho, Spencer Xu, Rafael Plana Simões

## Abstract

**Objective:** Biopsychosocial models recognize multiple determinants of post-trauma mental disorders, but their relative and interactive effects remain unclear. We quantified the independent contribution of traumatic event severity, preexisting vulnerability, social support, and coping capacity, and tested mediation pathways.

**Methods:** In a Brazilian clinical sample reporting traumatic or stressful events (N = 612), constructs were operationalized as composite scores and a dichotomous clinical outcome was derived from intake assessments. Logistic regression (n = 594) and structural equation modeling evaluated prediction and mediation.

**Results:** Vulnerability was the strongest risk factor (OR = 1.46, p < .001) and social support the main protective factor (OR = 0.60, p < .001). Traumatic event severity remained an independent predictor (OR = 1.39, p < .001), whereas coping capacity was not significant (OR = 0.94, p = .410). Discrimination was good (AUC = 0.80). Mediation indicated vulnerability reduced social support and coping capacity, with a significant indirect effect via social support.

**Conclusions:** Findings support a multifactorial model centered on a triad of vulnerability, social support, and traumatic exposure. Risk is shaped primarily by preexisting vulnerability and relational context, alongside a direct trauma effect, providing a clinically relevant framework for assessment and intervention.

## INTRODUCTION

Exposure to potentially traumatic events (PTEs) is common, with more than 70% of individuals reporting such experiences across the lifespan (Kessler et al., 2017). Yet most exposed individuals do not develop psychiatric disorders such as post-traumatic stress disorder (PTSD) (Bonanno & Mancini, 2012). This variability underscores a question in trauma research: why does a similar event result in disorder in some individuals but not in others?

Biopsychosocial models conceptualize trauma-related disorders as emerging from the interaction between event characteristics, preexisting vulnerabilities, and protective resources (Engel, 1977; Weisæth, 1998). Core determinants include trauma severity (Benjet et al., 2015), developmental and biological susceptibility (Yehuda & LeDoux, 2007), social support (Calhoun et al., 2022), and coping capacity (Badour & Feldner, 2013). Although these factors are independently associated with psychopathology, less is known about their relative predictive weight and interdependence within an integrated clinical model (Fink & Galea, 2015; Helpman et al., 2017).

Most studies examine these constructs separately or rely on bivariate associations, limiting understanding of how vulnerability and relational context jointly structure trauma outcomes. In particular, few investigations formally test both predictive discrimination and mediation pathways within a unified framework (Redican et al., 2021; Kerbage & Purper-Ouakil, 2024), especially in real-world clinical samples from low- and middle-income countries.

The present study aimed to test and refine an integrative model of post-traumatic mental disorder in a Brazilian clinical sample. Specifically, we sought to (1) quantify the independent predictive contribution of traumatic event severity, preexisting vulnerability, social support, and coping capacity, and (2) examine mediation pathways through which event and vulnerability may influence disorder via reductions in social support and coping capacity. By doing so, we propose a predictive and mechanistic framework with direct clinical relevance.

## METHOD

### Study Design and Sample

This analytical cross-sectional study included 905 individuals receiving psychological or psychiatric care, recruited by convenience sampling between October 2024 and July 2025 from Brazilian mental health services (primary care units, psychosocial care centers, hospitals, and private clinics). For predictive and mediation analyses, a subsample of 612 participants who self-reported a lifetime traumatic or stressful event at intake was analyzed. The study was approved by the Research Ethics Committee of the Centro Universitário de Ourinhos (CAAE: 78783724.0.0000.0332). All participants provided written informed consent prior to participation, in accordance with the Declaration of Helsinki.

### Measures and Data Preparation

Data were obtained through a structured intake interview comprising 213 variables. A theory-driven reduction procedure identified 28 indicators representing four core domains: Traumatic Event, Preexisting Vulnerability (biological and psychological), Social Support (contextual and relational factors), and Coping Capacity.

Variables were selected based on theoretical relevance, construct coverage, measurement structure (preference for categorical or scaled responses), data completeness, and non-redundancy. The final set ensured balanced representation across domains.

For each domain, composite scores (0–10) were created by numerically mapping categorical responses and aggregating standardized indicators. Scores were normalized to facilitate comparability.

The dichotomous outcome variable (DEVELOP_TRA) represented a pragmatic clinical proxy for mental disorder, derived from clinician assessment during intake combined with structured screening of the main complaint field. This measure reflects clinical classification rather than diagnosis established through standardized research interviews.

### Analytical Procedure

All analyses were conducted in R (version 4.3.2).

Descriptive statistics and Pearson correlations were calculated for the four domain scores.

To assess independent predictive effects, a binomial logistic regression model was fitted using complete cases (n = 594). Predictor variables included the four domain scores, and the outcome was DEVELOP_TRA. Results are presented as Odds Ratios (OR) with 95% confidence intervals. Model discrimination was evaluated using the Area Under the ROC Curve (AUC).

To examine mediation mechanisms, a dual mediation model was tested using Structural Equation Modeling (SEM). The model assessed whether the effects of Traumatic Event and Vulnerability on DEVELOP_TRA were mediated by Social Support and Coping Capacity. Model fit was evaluated using CFI, TLI, RMSEA, and SRMR indices. Indirect effects were tested for statistical significance.

## RESULTS

### Descriptive statistics of the sample and component scores

The four theoretical pillars were operationalized as normalized continuous scores (0–10). Descriptive statistics for the analytical sample (n = 612) are presented in Table 1. All scores showed adequate variability for inferential analyses. Skewness and kurtosis values were within acceptable limits for parametric modeling (absolute values < 2), although Vulnerability showed moderate positive skewness (1.08).

**Table 1.**
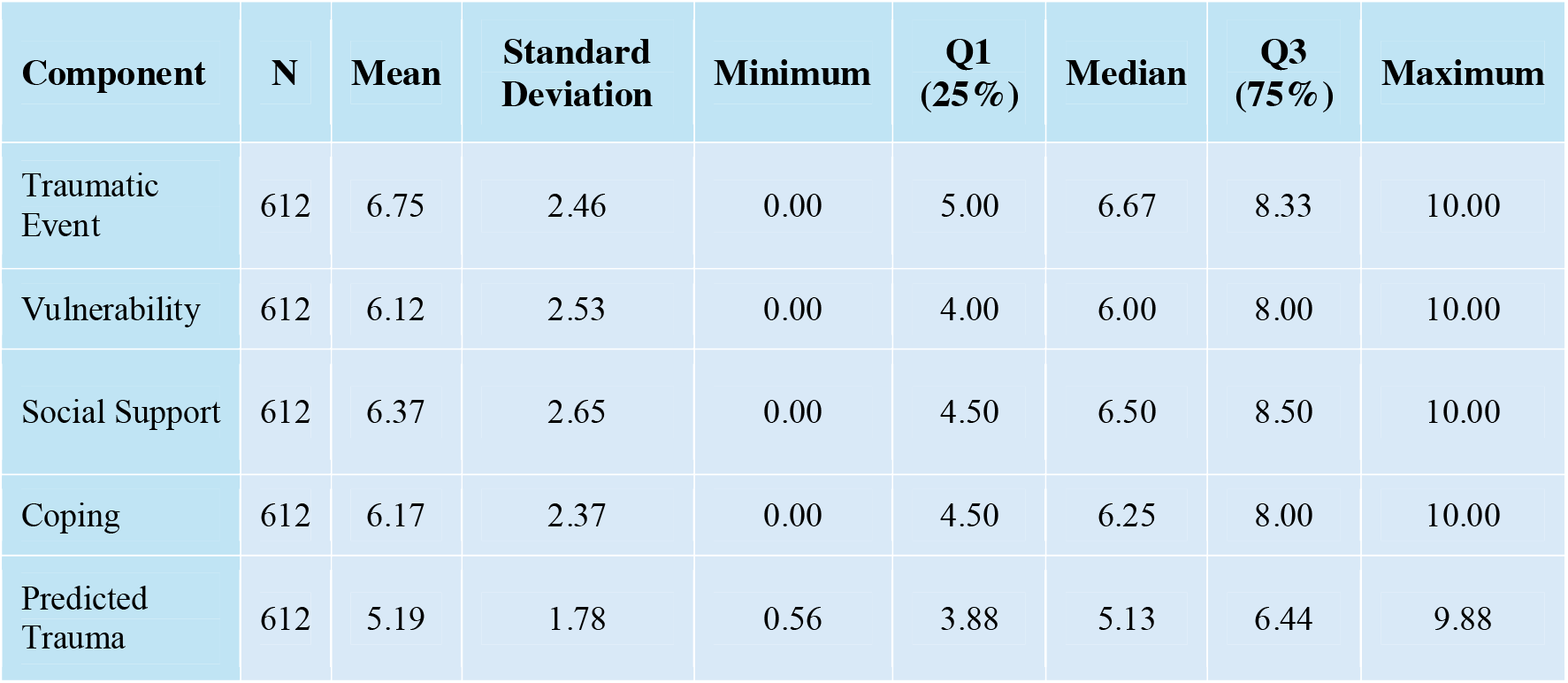
Descriptive Statistics of Component Scores (Real Data, n=612)

| Component | N | Mean | Standard Deviation | Minimum | Q1 (25%) | Median | Q3 (75%) | Maximum |
| --- | --- | --- | --- | --- | --- | --- | --- | --- |
| Traumatic Event | 612 | 6.75 | 2.46 | 0.00 | 5.00 | 6.67 | 8.33 | 10.00 |
| Vulnerability | 612 | 6.12 | 2.53 | 0.00 | 4.00 | 6.00 | 8.00 | 10.00 |
| Social Support | 612 | 6.37 | 2.65 | 0.00 | 4.50 | 6.50 | 8.50 | 10.00 |
| Coping | 612 | 6.17 | 2.37 | 0.00 | 4.50 | 6.25 | 8.00 | 10.00 |
| Predicted Trauma | 612 | 5.19 | 1.78 | 0.56 | 3.88 | 5.13 | 6.44 | 9.88 |

### Bivariate analyses and preliminary framework validation

Pearson correlations were calculated between the four pillar scores and the composite Predicted Trauma Score. As shown in Table 2, all associations were statistically significant (p < .001) and consistent with theoretical expectations.

**Table 2.** Bivariate correlations between theoretical pillars and the predicted trauma score (n = 612).

| Pillar | Correlation (r) | 95% CI | p-value |
| --- | --- | --- | --- |
| Social Support | -0.62 | [-0.67, -0.57] | < .001 |
| Traumatic Event | +0.63 | [+0.58, +0.67] | < .001 |
| Vulnerability | +0.59 | [+0.54, +0.64] | < .001 |
| Coping Capacity | -0.45 | [-0.51, -0.39] | < .001 |

Note: CI = Confidence Interval. All correlations are two-tailed.

Traumatic Event showed the strongest positive correlation (r = .63), followed by Preexisting Vulnerability (r = .59). Social Support demonstrated a strong negative association (r = −.62), and Coping Capacity showed a moderate negative correlation (r = −.45).

Because the Predicted Trauma score was constructed from the same theoretical pillars used as predictors in subsequent analyses, these bivariate correlations reflect internal coherence rather than independent validation. Their function is to demonstrate directional consistency among model components.

As expected, each pillar was significantly associated with the overall trauma score in the hypothesized direction. However, these bivariate associations do not account for shared variance among predictors and therefore cannot determine their unique contribution. This issue is addressed in the multivariate analyses.

### Optimized predictive model of mental disorder development

To examine the independent predictive contribution of each pillar, a binomial logistic regression model was fitted using DEVELOP_TRA (presence/absence of mental disorder) as the outcome. This variable was derived from the severity and chronicity of the chief complaint according to a standardized clinical classification protocol aligned with DSM-5-TR criteria. After excluding cases with missing data, the final analytical sample comprised 594 participants.

As shown in Table 3, the model revealed a clear hierarchy among predictors. Preexisting Vulnerability emerged as the strongest risk factor (β = 0.38, *p* < .001; OR = 1.46). Social Support was the primary protective factor (β = −0.51, *p* < .001; OR = 0.60). Traumatic Event severity remained a significant independent predictor (β = 0.33, *p* < .001; OR = 1.39). In contrast, Coping Capacity did not retain statistical significance in the multivariate model (β = −0.06, *p* = .410; OR = 0.94).

**Table 3.** Results of the logistic regression for the prediction of mental disorder development (n = 594).

| Predictor | Coefficient ($\beta$) | Std. Error | Odds Ratio (OR) | 95% CI for OR | p-value |
| --- | --- | --- | --- | --- | --- |
| (Intercept) | -4.49 | 0.63 | 0.01 | [0.00, 0.02] | < .001 |
| Traumatic Event | 0.33 | 0.07 | 1.39 | [1.22, 1.58] | < .001 |
| Vulnerability | 0.38 | 0.06 | 1.46 | [1.30, 1.64] | < .001 |
| Social Support | -0.51 | 0.06 | 0.60 | [0.53, 0.68] | < .001 |
| Coping Capacity | -0.06 | 0.07 | 0.94 | [0.82, 1.08] | .410 |

Note: The outcome variable is DEVELOP\_TRA (1 = Yes, 0 = No). CI = Confidence Interval.

The model demonstrated good discriminatory performance (AUC = 0.80, 95% CI: 0.76–0.83).

### Analysis of underlying mediation mechanisms

The structural model showed acceptable fit: χ^2^(1) = 4.85, *p* = .028; CFI = 0.980; TLI = 0.929; RMSEA = 0.075 (90% CI: 0.018–0.149); SRMR = 0.031.

Preexisting Vulnerability was significantly associated with lower Social Support (β = −0.40, *p* < .001), which in turn predicted higher probability of disorder (β = −0.53, *p* < .001). The indirect effect of Vulnerability via Social Support was significant (β = 0.21, *p* < .001), while the direct effect of Vulnerability remained significant (β = 0.31, *p* < .001). Vulnerability was also associated with lower Coping Capacity (β = −0.22, *p* < .001).

The pathway from Traumatic Event to Social Support was marginal (β = −0.09, *p* = .059). However, the direct effect of Traumatic Event on the outcome remained significant in the mediation model (β = 0.18, *p* = .002). Pathways involving Coping Capacity as mediator were not statistically significant (β = −0.02, *p* = .724).

Although CFI and SRMR indicated good fit, RMSEA suggests cautious interpretation, consistent with cross-sectional mediation models.

**Figure 1.**
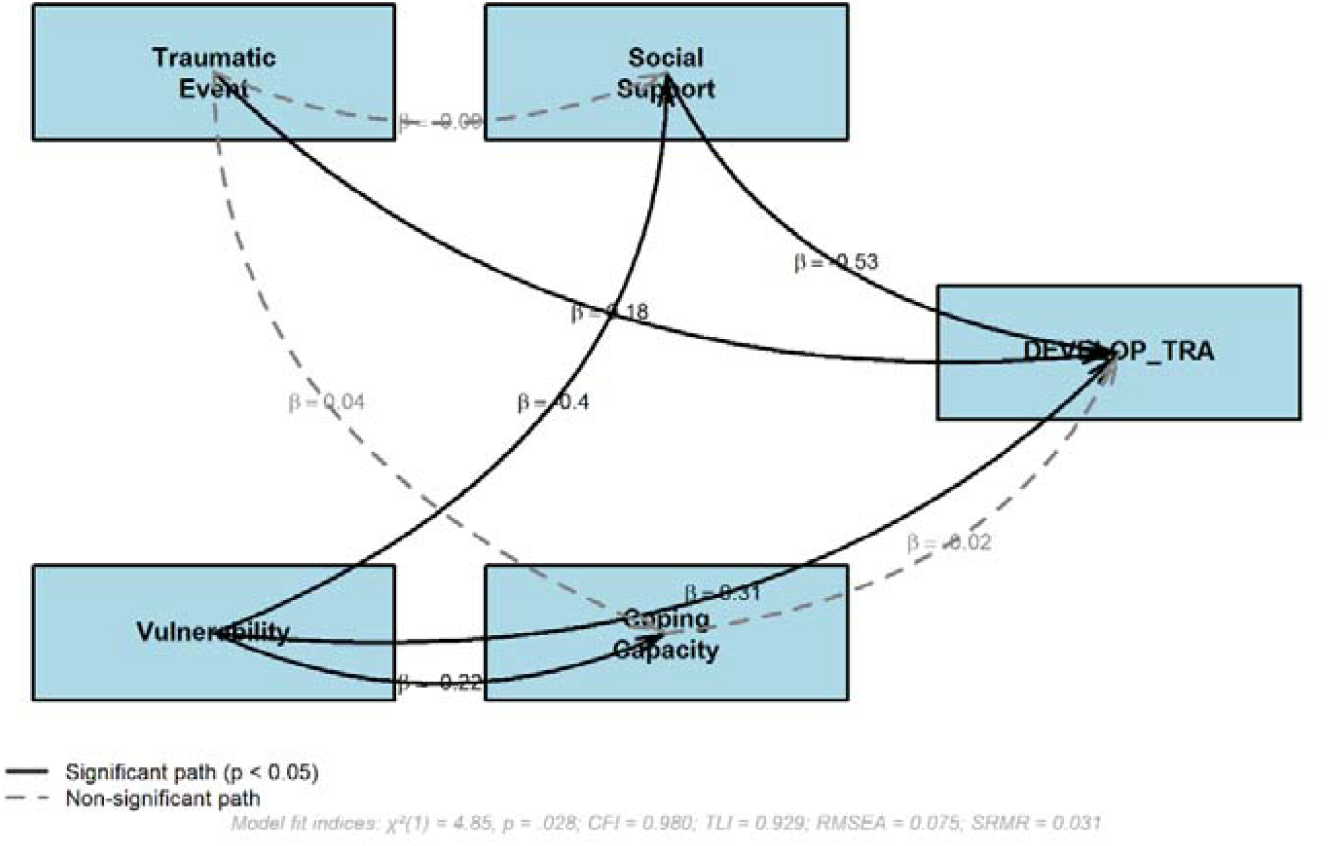
Path diagram of the dual mediation model with standardized coefficients (β). *Note:* Solid lines indicate significant paths (*p* < .05); dashed lines indicate non-significant paths. Coefficients are standardized. The outcome variable is DEVELOP_TRA.

## DISCUSSION

In our broader clinical dataset (N = 905; 213 variables), 612 participants (67.6%) reported having experienced a traumatic or stressful event during their lifetime. This high prevalence reflects the well-documented co-occurrence between trauma exposure and mental health complaints in clinical populations (Turner & Lloyd, 1995; Morales-Toro et al., 2019). However, exposure alone is insufficient to determine outcome.

Epidemiological research consistently demonstrates that traumatic events are common, yet only a subset of exposed individuals develop psychiatric disorders (Kessler et al., 2017). As noted by McQuaid et al. (2001), trauma is a necessary criterion for PTSD, but its occurrence is more frequently associated with other disorders, particularly major depressive disorder. Thus, the presence of trauma does not function as a deterministic predictor of psychopathology.

Conceptually, traumatic experiences vary both in objective characteristics and subjective appraisal. While certain events are widely recognized as severe (e.g., interpersonal violence, bereavement), other experiences may acquire traumatic significance depending on personal meaning and perceived threat. This aligns with stress appraisal frameworks that emphasize the relational evaluation between the individual and the environment (Lazarus, 1984). In clinical contexts, qualitative reports often reveal this heterogeneity, encompassing both conventionally defined traumas and events with strong subjective impact.

The high prevalence of self-reported trauma in our sample therefore reinforces a paradox: exposure is frequent, yet disorder is not universal. The critical question becomes not whether trauma occurred, but under which biopsychosocial conditions exposure becomes associated with clinically significant mental disorder.

Extensive literature establishes that the nature and severity of traumatic events are associated with psychopathological outcomes. Adverse childhood experiences show a clear dose–response relationship with adult mental disorders (McKay et al., 2022), and early adversity is consistently linked to increased risk of common mental disorders and suicidality (Sahle et al., 2022). In adulthood, acute stressors such as violence, injury, and bereavement further elevate psychiatric risk (Chen et al., 2024), and large-scale stressors such as natural disasters can precipitate clinically significant disorders depending on exposure severity and contextual factors (Saeed & Gargano, 2022).

Nevertheless, exposure alone remains an insufficient predictor. Contemporary biopsychosocial models emphasize that outcomes emerge from the interaction between stressor characteristics and individual resources (Matheson et al., 2020). The critical question shifts from whether trauma occurred to how vulnerability and protective factors structure post-trauma trajectories.

Our findings support this multifactorial perspective while refining its internal organization. Although bivariate analyses indicated significant associations across all four pillars, multivariate modeling revealed a structured hierarchy. Preexisting vulnerability emerged as the strongest independent predictor (OR = 1.46, *p* < .001), even after controlling for trauma severity and social support. This contrasts with earlier syntheses suggesting stronger effects for peri- and post-traumatic factors (Brewin et al., 2000), indicating that in clinical samples, prior susceptibilities may exert greater pathogenic influence when modeled simultaneously.

Social support functioned as the primary protective factor (OR = 0.60, *p* < .001), reinforcing the centrality of relational context. Within this domain, contemporary distinctions between objective network availability and subjective perceived connection warrant attention. In our operationalization, proxy indicators captured both structural and perceived aspects of support, allowing examination of their relative contributions within the integrated model.

Taken together, these findings support a hierarchical relational vulnerability framework in which preexisting susceptibility and relational context organize the impact of traumatic exposure, shaping whether adversity culminates in clinically significant disorder.

This exploratory analysis yielded two main observations. First, objective network availability and subjective perceived support were empirically distinct (r ≈ 0.20), supporting their conceptual separation. Second, objective isolation emerged as a marginally stronger predictor than subjective loneliness, although neither retained independent significance after adjustment for trauma severity and vulnerability. In this clinical sample, the absence of accessible support structures may represent a more immediate vulnerability than perceived disconnection alone.

The traumatic event remained a significant independent predictor (OR = 1.39, *p* < .001), consistent with evidence identifying trauma as a transdiagnostic risk factor with heterogeneous effect sizes (Hogg et al., 2023). In the integrated model, event severity exerted a concurrent and direct effect, while also operating within a network shaped by preexisting vulnerability (OR = 1.46) and relational context (OR = 0.60). This supports a framework in which trauma impact is neither purely deterministic nor solely indirect, but structurally embedded within prior susceptibility and social resources.

Coping capacity did not retain an independent protective effect in the multivariate model (OR = 0.94, *p* = .410). Mediation analyses indicated that vulnerability was significantly associated with reduced coping (β = -0.22, *p* < .001), suggesting that coping may function as a contingent resource influenced by more foundational individual and relational factors rather than as a primary resilience pillar.

The mediation structure clarified the mechanisms linking vulnerability to outcome. Vulnerability predicted reduced social support (β = -0.40, *p* < .001) and reduced coping capacity, with a significant indirect pathway to disorder through social support (β = 0.21, *p* < .001). Importantly, the traumatic event retained a direct effect in the full model (β = 0.18, *p* = .002), reinforcing its independent contribution.

The model demonstrated good discriminatory performance (AUC = 0.80), supporting its clinical applicability in identifying individuals at elevated risk following trauma exposure.

### Clinical Implications

These findings translate into clear, nuanced priorities for clinical practice and public health, advocating for an integrated focus that addresses both the significant impact of the traumatic event and the individual’s biographical and relational context. Consequently, clinical evaluations must systematically assess three core elements: the nature and subjective severity of the index traumatic event; preexisting vulnerabilities (developmental history, past trauma);and a meticulous mapping of current social support networks. This tripartite biopsychosocial map has proven highly predictive and must form the cornerstone of clinical case formulation.

Building on this assessment, therapeutic strategies should be strategically sequenced with dual initial priorities. First, validate and address the direct impact of the traumatic event through stabilization and trauma-informed care. Concurrently and with equal emphasis, intervention should focus on actively rebuilding and strengthening the patient’s accessible social support system, our strongest identified protective factor. This directly targets the key mediating pathway where vulnerability erodes support (Vulnerability → Reduced Support → Disorder). Practical avenues include facilitating access to support groups, employing family or couple therapy, and fostering community integration.

Establishing this stabilized and fortified foundation creates the secure context from which to address, in a subsequent stage, the underlying vulnerabilities through appropriate trauma-focused or psychodynamic modalities. Within this strengthened environment, the role of coping skills training can be optimally framed. Our results suggest that isolated coping training may be less effective; therefore, these strategies should be introduced not as a standalone solution, but as an integrated set of tools to help individuals manage distress within their now more robust relational network, thereby maximizing their modifiable potential.

Finally, the implications extend beyond the clinic. At a public health level, these results underscore the preventive power of social connectedness. Policies and programs designed to bolster community cohesion and social infrastructure represent a highly efficient universal strategy to build population-wide resilience, potentially buffering the impact of adverse events and mitigating the effects of preexisting vulnerabilities across communities.

### Limitations

This study has limitations that must be considered when interpreting its findings. First, the cross-sectional design precludes definitive causal inferences, although our mediation model proposes plausible pathways, longitudinal data are required to firmly establish the directionality and temporal sequence of these effects.

Additionally, the outcome variable should be interpreted as a pragmatic clinical indicator rather than a gold-standard diagnostic measure, as it was not derived from structured diagnostic interviews such as SCID or MINI. While this approach enhances ecological validity, it may introduce classification variability.

Second, while substantial, our sample was a clinical convenience sample recruited from Brazilian mental health services. This may affect the generalizability of the results to non-clinical populations, other healthcare systems, and importantly, to different cultural contexts where the expression of trauma, social support, and coping may vary.

Third, the operationalization of the primary outcome relied on diagnoses made by treating clinicians during routine intake interviews, rather than assessments conducted with research-standardized diagnostic instruments (e.g., SCID, MINI). Although this method reflects real-world clinical practice and benefits from professional judgment, it may lack the consistency, specificity, and reliability of controlled diagnostic protocols used in dedicated research settings.

Finally, the non-significant independent role of Coping Capacity in our multivariate model warrants careful interpretation. This finding may reflect the specific measurement of coping in this study, the high interdependence of coping with vulnerability and social support in our clinical sample—as evidenced by the significant path from Vulnerability to lower Coping Capacity in the mediation model—or a true contextual effect. Replication in independent samples with diverse measurements of coping is needed to determine whether its role is consistently secondary or mediated in integrated models of post-trauma outcome.

### Future Directions

Longitudinal studies are needed to validate the proposed mediation pathways and establish temporal sequencing. Replication in diverse cultural and clinical populations is required to assess generalizability. Further research should refine measurement of social support components and clarify the contextual role of coping within integrated models of trauma outcome.

## CONCLUSION

This study advances the understanding of trauma-related mental disorders by testing an integrated biopsychosocial model from predictive and mechanistic perspectives. The findings support a multifactorial etiology while identifying a structured hierarchy among determinants.

Risk of disorder was best explained by a core triad: preexisting vulnerability as the primary risk factor, social support as the principal protective resource, and traumatic event severity as a significant independent predictor with both direct and concurrent effects. Coping capacity did not function as an independent protective pillar in the integrated model,suggesting that its influence may depend on more foundational individual and relational conditions.

The mediation analysis clarified key mechanisms. Preexisting vulnerability exerted its effect both directly and indirectly by reducing social support and coping capacity, with the indirect pathway through social support being particularly robust. These findings underscore the centrality of relational context in post-trauma outcomes and support interventions that strengthen support networks alongside addressing trauma exposure and underlying vulnerability.

Given the cross-sectional design, causal inference remains limited. Longitudinal studies are needed to validate the proposed pathways and further clarify the contextual role of coping. The weighted triadic framework presented here offers a practical structure for clinical risk assessment and intervention planning.

## Data Availability

All data produced in the present study are available upon reasonable request to the authors

## ACKNOWLEDGMENTS

The authors gratefully acknowledge the financial support provided by the Coordenação de Aperfeiçoamento de Pessoal de Nível Superior (CAPES, Brazil) and the Institute of Collaborative Innovation (ICI), University of Macau. We also thank the participants who generously contributed their time and experiences to this study.

We are particularly grateful to the students and staff of Centro Universitário de Ourinhos/SP and Centro de Ensino “Eliska Sedlák” of Santa Casa de Ourinhos/SP for their collaboration during data collection and logistical support. We additionally acknowledge the support of Universidad de Los Lagos (Puerto Aysén) for institutional cooperation.

The authors thank colleagues and collaborators who provided valuable assistance with language revision, manuscript preparation, and methodological discussions throughout the research process. Their contributions were essential to the development and refinement of this study.

